# Post-mortem Nasopharyngeal Microbiome Analysis of Zambian Infants with and without Respiratory Syncytial Virus Disease: A Nested Case Control Study

**DOI:** 10.1101/2022.12.23.22283745

**Authors:** Jessica McClintock, Aubrey R. Odom-Mabey, Nitsueh Kebere, Arshad Ismail, Lawrence Mwananyanda, Christopher J. Gill, William B. MacLeod, Rachel C. Pieciak, Rotem Lapidot, W. Evan Johnson

**Affiliations:** Division of Infectious Disease, Center for Data Science, Rutgers New Jersey Medical School, Newark, NJ, 07103, USA; Bioinformatics Program, Boston University, Boston, MA, 02118, USA; Sequencing Core Facility, National Institute for Communicable Diseases of the National Health Laboratory Service, 2131 Johannesburg, South Africa; Department of Biochemistry and Microbiology, University of Venda, Thohoyandou 0950, South Africa; Department of Global Health, Boston University School of Public Health, Boston, MA, 02118, USA; Pediatric Infectious Diseases, Boston Medical Center, Boston, MA, 02118, USA; Pediatrics, Boston University School of Medicine, Boston, MA, 02118, USA

## Abstract

**Background:** Respiratory Syncytial Virus (RSV) is the most common cause of bronchiolitis and lower respiratory tract infections in children in their first year of life, disproportionately affecting infants in developing countries. Previous studies have found that the nasopharyngeal microbiome of infants with RSV infection has specific characteristics that correlate with disease severity, including lower biodiversity, perturbations of the microbiota and differences in relative abundance. These studies have focused on infants seen in clinical or hospital settings, predominantly in developed countries.

**Methods:** We conducted a nested case control study within a random sample of 50 deceased RSV+ infants with age at death ranging from 4 days to 6 months and 50 matched deceased RSV-infants who were all previously enrolled in the Zambia Pertussis and RSV Infant Mortality Estimation (ZPRIME) study. All infants died within the community or within 48 hours of facility admittance. As part of the ZPRIME study procedures, all decedents underwent one-time, post-mortem nasopharyngeal sampling. The current analysis explored the differences between the nasopharyngeal microbiome profiles of RSV+ and RSV-decedents using 16S ribosomal DNA sequencing.

**Results:** We found that *Moraxella* was more abundant in the nasopharyngeal microbiome of RSV+ decedents than in RSV-decedents. Additionally, *Gemella* and *Staphylococcus* were less abundant in RSV+ decedents than in RSV-decedents.

**Conclusion:** These results support previously reported findings of the association between the nasopharyngeal microbiome and RSV and suggest that changes in the abundance of these microbes are likely specific to RSV and may correlate with mortality associated with the disease.

## INTRODUCTION

Respiratory syncytial virus (RSV) infection is the most common cause of bronchiolitis and pneumonia in children in the first year of life.^1,2^ Symptoms of RSV range from mild upper respiratory infections to more severe infections such as bronchiolitis and pneumonia.^3,4^ RSV has been established as the leading global cause of lower respiratory tract infections in infants, with an etiological fraction for pneumonia three times higher than the next highest named pathogen.^2^ RSV continues to be a major health concern in both developed and developing nations.^5,6^ The developing world experiences the highest burden of RSV infection, accounting for 99% of world-wide deaths from RSV-related lower respiratory tract infections in children under five.^3^ We recently reported that in low-income countries, roughly two-thirds of infant RSV deaths occurred in the community.^7^

Several studies have looked at associations between the nasopharyngeal (NP) microbiome profiles of infants and RSV disease severity. Previous studies have shown that the NP microbiome in RSV-infected infants has characteristics that changed with severity of disease.^2,8–13^ For instance, an overabundance of *Achromobacter, Haemophilus, Moraxella*, and *Streptococcus* microbes and a loss of *Corynebacterium, Staphylococcus*, and *Veillonella* have been associated with infants with more severe RSV.^11–13^ Studies have also shown that antibiotic use leads to decreased alpha diversity and dysbiosis in the NP microbiome.^14,15^

RSV infection may include indirect effects mediated by its impact on other members of the respiratory ecosystem.^16–18^ These effects seem to occur with bacteria such as *Haemophilus influenzae* and *Streptococcus pneumoniae*, suggesting that the impact of RSV on bacterial pathogenesis is relatively specific.^19,20^

Data from studies outside of high-income countries is very limited, and yet most RSV-associated mortality is concentrated in low- and middle-income countries.^6^ Until recently, the burden of RSV in low- and middle-income countries was estimated with hospital-based surveillance data used to model RSV prevalence among community deaths. The Zambia Pertussis and RSV Infant Mortality Estimation (ZPRIME) study was a systematic, post-mortem surveillance study designed to address this knowledge gap by directly measuring facility and community deaths in Lusaka, Zambia.^7^ The ZPRIME study found that RSV caused 2.8% of all infant deaths and 4.7% of all community deaths.

Our current analysis examines the NP microbiomes of a subset of RSV+ and RSV-deceased infants enrolled in the ZPRIME study. 16S ribosomal DNA sequencing was conducted on post-mortem infant NP samples with the goal of characterizing and comparing the NP microbiome profiles of RSV- and RSV+ decedents. Our study focuses on community deaths in a low-income county to address gaps from previous studies.

## METHODS

### Study Population and Study Design

We identified a nested cross-sectional sample of RSV+ and RSV-deceased infants from the ZPRIME study.^7^ ZPRIME aimed to determine the proportion of infant deaths that could be attributed to *Bordetella pertussis* and RSV infections. Decedents were between four days and six months old and enrolled in the ZPRIME study within 48 hours of death. We selected a subset of decedents for our analysis who were RSV+ with a PCR Ct<35. Demographic and clinical data, including sex, age at death, mother’s human immunodeficiency virus (HIV) status, location of death, and cause of death, were collected for each decedent (Table 1). All decedents underwent one-time, post-mortem NP sampling. NP samples were tested for RSV by reverse transcriptase quantitative PCR following the testing protocol developed by the Centers for Disease Control and Surveillance.^21^

**Table 1.** Characteristics of deceased RSV+ and RSV-infants. RSV+ infants were selected through random sampling and were matched to RSV-infants by age at death and date of death. Chi-squared tests were performed for each group. Note that the infant’s sex was not reported in all cases.

| Characteristics of RSV+ and RSV- Decedents |  |  |  |  |
| --- | --- | --- | --- | --- |
|  | RSV+ Infants | RSV- Infants | All Subjects | p-value |
| *In study (n)* | 50 | 50 | 100 |  |
| **Sex** |  |  |  | 0.128 |
| *Males % (n)* | 46% (23/50) | 56% (28/50) | 51% (51/100) |  |
| *Females % (n)* | 38% (19/50) | 40% (20/50) | 39% (39/100) |  |
| *Unknown Sex % (n)* | 16% (8/50) | 4% (2/50) | 10% (10/100) |  |
| **Age** |  |  |  | 0.369 |
| *Median Age (days)* | 41 | 39 | 40.5 |  |
| *4-28 days % (n)* | 38% (19/50) | 38% (19/50) | 38% (38/100) |  |
| *29-61 days % (n)* | 30% (15/50) | 28% (14/50) | 29% (29/100) |  |
| *62-152 days % (n)* | 24% (12/50) | 26% (13/50) | 25% (25/100) |  |
| *153-182 days % (n)* | 8% (4/50) | 8% (4/50) | 8% (8/100) |  |
| **Mother's HIV Status** |  |  |  | 0.269 |
| *HIV Exposed % (n)* | 4% (2/50) | 12% (6/50) | 8% (8/100) |  |
| *HIV Unexposed % (n)* | 96% (48/50) | 88% (44/50) | 92% (92/100) |  |
| **Location of Death** |  |  |  | 1.000 |
| *Early Facility Death % (n)* | 14% (7/50) | 16% (8/50) | 15% (15/100) |  |
| *Community Death % (n)* | 86% (43/50) | 84% (42/50) | 85% (85/100) |  |
| **Cause of Death** |  |  |  | <0.001 |
| *Respiratory % (n)* | 72% (36/50) | 26% (13/50) | 49% (49/100) |  |
| *Non-Respiratory % (n)* | 22% (11/50) | 64% (32/50) | 43% (43/100) |  |
| *Unsure % (n)* | 6% (3/50) | 10% (5/50) | 8% (8/100) |  |

We selected 50 RSV+ decedents via random sampling and matched those to 50 RSV-decedents. Due to age- and seasonality-related variations in RSV impact, we matched decedents by age at death (4–28 days (0+ months), 29–61 days (1+ month), 62–152 days (2+ months) and 152–182 days (4+ months)) and date of death (+/- 1.5 months) (Table 1).

Additional details about our study population are available in Text, Supplemental Digital Content 1, as well as information about sample collection, processing, and storage, 16S ribosomal rDNA amplification and sequencing, and data processing.

### Statistical Analysis

We analyzed microbe counts at both the genus and species levels, which is possible using PathoScope2.0.^22–24^ We performed chi-squared tests to determine if the population characteristics of our RSV+ and RSV-groups were evenly distributed. We calculated relative abundances based on raw count data for the taxon grouped by disease state and constructed bar plots, boxplots, and heatmaps comparing microbe prevalence across the RSV+ and RSV-conditions.^25,26^ Samples in the bar chart are ordered by the top 4 most abundant samples found in RSV+ samples to enhance visual organization. Differential abundance analysis was conducted based on relative abundance between genera using a nonparametric Wilcoxon rank sum test.^27–29^ To correct for multiple testing, we used Benjamini-Hochberg p-value adjustments.^30^ Significant results, defined as p<0.05 and adjusted p<0.25, were visualized with boxplots.

To analyze alpha diversity, we computed Shannon and Simpson indices and visualized each index as a boxplot.^31,32^ We compared indices across RSV state using Wilcoxon rank sum test as the data was not normally distributed, as determined by the Shapiro-Wilk test for normality.^27–29,33–35^

To analyze beta diversity, we computed the Bray–Curtis dissimilarity indices to observe differences in microbial composition between the samples.^36^ The results were visualized using non-metric dimensional scaling (NMDS) and principal coordinate analysis (PCoA).^26,37,38^ We performed NMDS with 20 stress test runs. Stress values are reported to indicate how well the model fits, where a lower stress value indicates a better model fit. Atchison distances were also computed and visualized via PCoA.^39^ Ellipses on the NMDS and PCoA plots represent a 95% confidence level for a multivariate t-distribution. PERMANOVA was performed to determine if the centroids and dispersion was statistically significant between RSV states.^40^

We analyzed pathway abundances via a Wilcoxon rank sum test with Benjamini-Hochberg corrections to determine if there was a difference in inferred pathway abundances between RSV+ and RSV-samples.^27–30,41–43^ Significant results have an adjusted p<0.05.

## RESULTS

### Data Filtering

Due to the heavy presence of taxa with relatively low abundances in the raw data, taxa with average relative abundances <0.1% were filtered from the dataset and regrouped as a single “Other” category. This reduced our identified taxa assignments from 632 genera encompassing 1042 species to 36 genera encompassing 54 species, not including the “Other” grouping. We used these data for all analyses expect for visualization of relative abundance, which used a data set including genera with average relative abundances <1%. This smaller group consisted of 15 genera and 1 “Other” category.

### Baseline characteristics of the study population

Table 1 displays the baseline characteristics for the 100 decedents. The groups were roughly even across sex (51% male, 39% female, 10% unknown). The median age of death was 40.5 days. The majority (85%) of decedents were brought in dead from the community. A small proportion of deceased infants in both groups were known to be born to mothers infected with HIV (8%), although the final HIV status of the decedents themselves remains unknown. Among all decedents, there were 49 respiratory deaths, 43 non-respiratory deaths and 8 inconclusive deaths. As expected, most RSV+ deaths were respiratory (72%) and most RSV-deaths were non-respiratory (64%) in origin.

Chi-squared tests were performed to determine if the population characteristics were evenly distributed between the RSV+ and matched RSV-decedents. No significant difference was found for sex (p=0.128), age (p=0.369), HIV exposure status (p=0.269), or location of death (p=1.000) indicating an even decedent distribution among groups (Table 1). The cause of death (respiratory vs non-respiratory) differed significantly between RSV+ and RSV-deceased infants (p<0.001).

### Relative Abundance

To compare the relative abundance of microbial taxa across RSV+ and RSV-samples, we generated a stacked bar plot composed of all genera with relative abundances greater than 1% (Figure 1). Figure 1 shows a total of 16 genera across all the samples with varying abundance levels. Each column represents the composite NP microbiome for a single participant and are ordered based on the abundance of the top 4 RSV+ microbes. When averaged across samples, the five most abundant microbes in RSV+ samples were *Streptococcus* (22.2%), *Haemophilus* (19.7%), *Moraxella* (11.0%), *Klebsiella* (10.2%), and *Corynebacterium* (6.8%). The five most abundant microbes in RSV-samples were *Streptococcus* (29.0%), *Staphylococcus* (13.5%), *Escherichia* (8.5%), *Klebsiella* (6.4%) and *Haemophilus* (5.7%).

**Figure 1.**
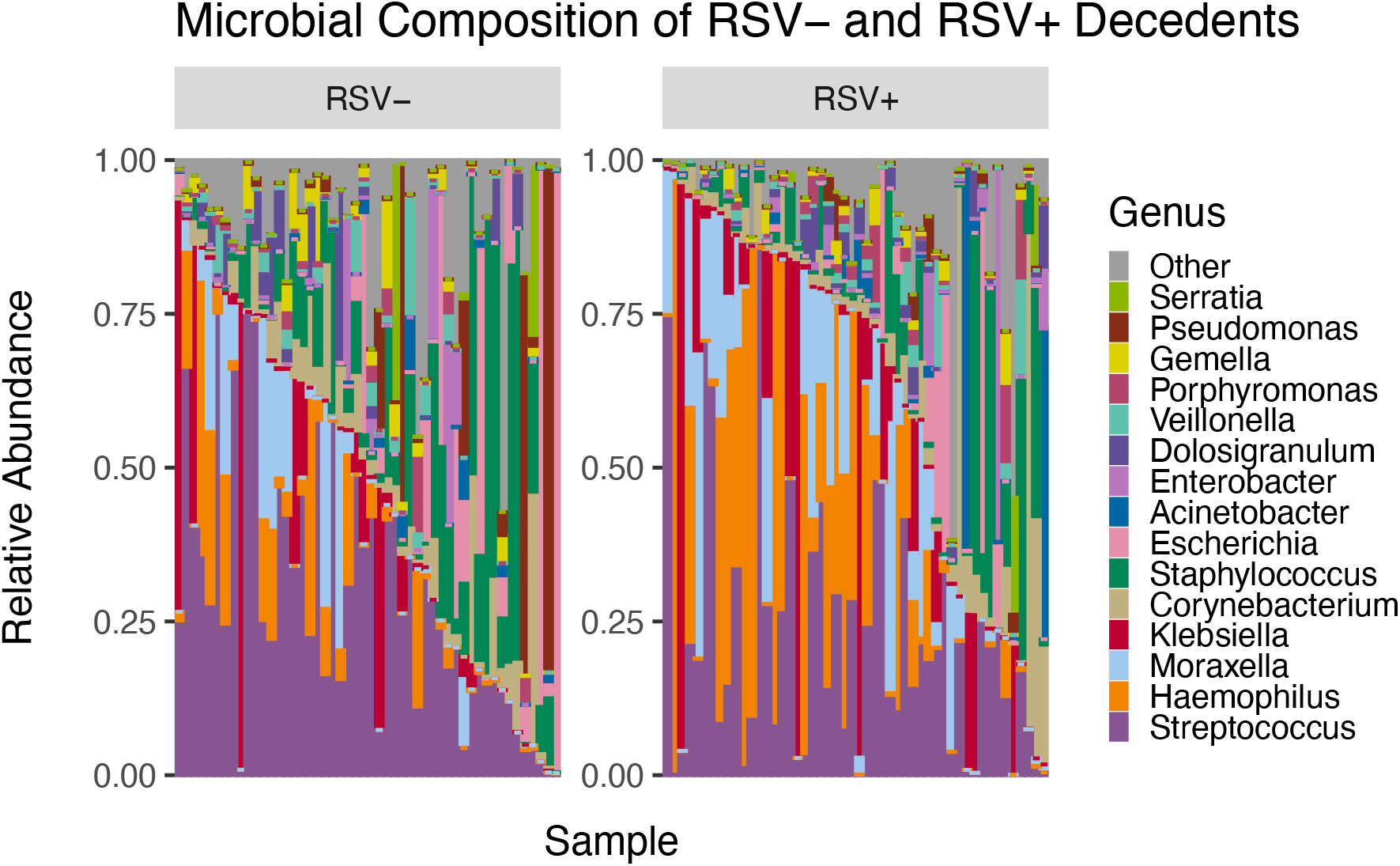
Microbial Composition of RSV- and RSV+ samples showing microbe diversity within each sample. On average, the five most abundant microbes in RSV+ samples were *Streptococcus* (22.2%), *Haemophilus* (19.7%), *Moraxella* (11.0%), *Klebsiella* (10.2%), and *Corynebacterium* (6.8%). The five most abundant microbes in RSV-samples were *Streptococcus* (29.0%), *Staphylococcus* (13.5%), *Escherichia* (8.5%), *Klebsiella* (6.4%) and *Haemophilus* (5.7%).

### Alpha Diversity

#### Shannon and Simpson Index

Shannon and Simpson indices were calculated and a Shapiro-Wilk test was performed for each variable (Figure 2A). The Shannon indices for genera did not follow a normal distribution (p=0.985 for RSV+ and p=0.860 for RSV-), whereas the Simpson indices did follow a normal distribution (p=0.002 for RSV+ and p<0.001 for RSV-). A Wilcoxon rank sum test of significance was performed to determine significance of the differences in the Shannon and Simpson indices. We observed no significant difference in the alpha diversity between the RSV+ and RSV-decedents for genera (p=0.621 for Observed, p=0.287 for Shannon and p=0.206 for Simpson). No significant alpha diversity differences were found when comparing species level taxa in the same manner (p=0.790 for Observed, 0.173 for Shannon, 0.120 for Simpson) (Figure 2B).

**Figure 2.**
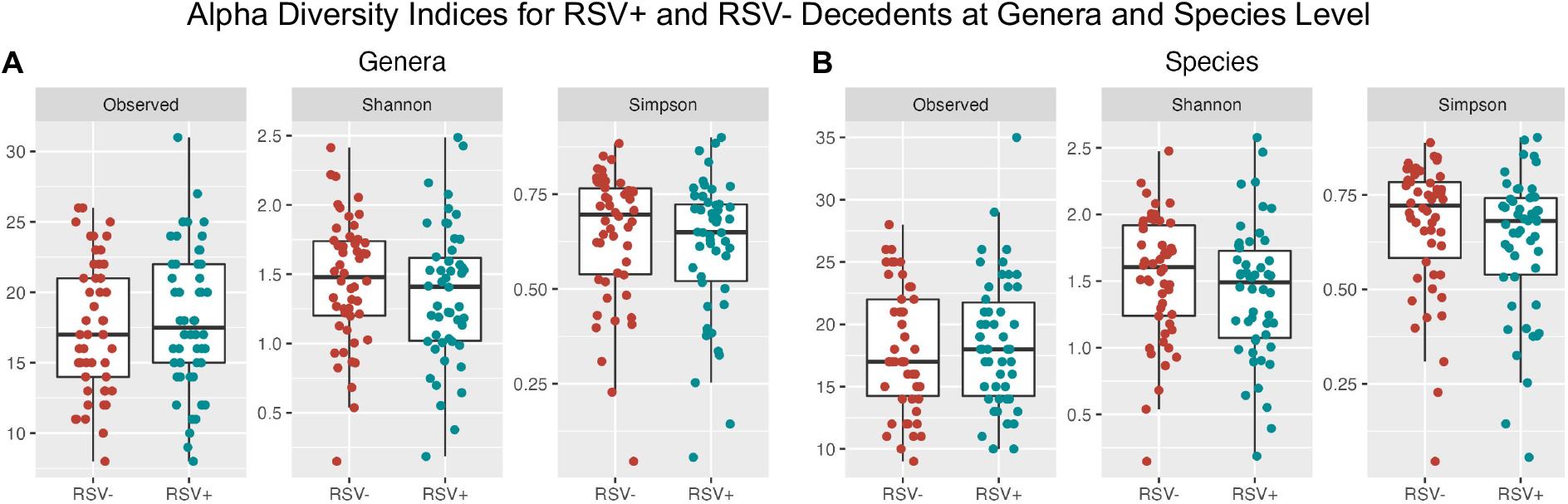
Alpha diversity for RSV+ and RSV-Zambian Decedents. No significant differences were found when conducting Wilcoxon rank sum test for **A)** genera (p=0.621 for Observed, 0.287 for Shannon, and 0.206 for Simpson) or **B)** species (p=0.790, 0.173, 0.120 respectively).

### Beta Diversity

#### Bray-Curtis Dissimilarity Index

We computed Bray-Curtis dissimilarity indices and visualized the results using NMDS and PCoA (Figure 3). Using PERMANOVA, we found that there was a significant difference in the centroids and dispersion between RSV groups (p<0.001).

**Figure 3.**
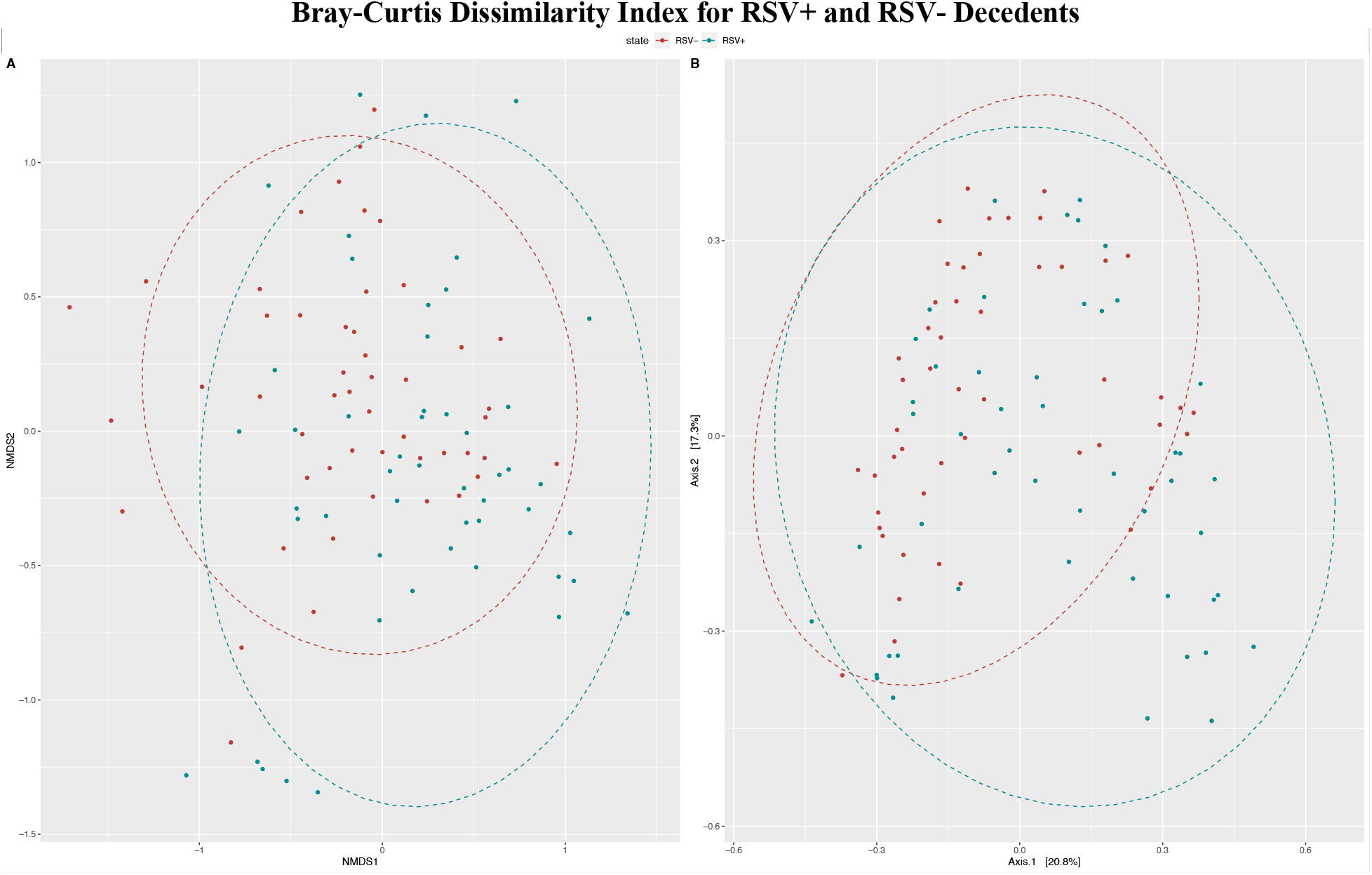
**A)** NMDS displaying the Bray-Curtis dissimilarity index for genera comparison between RSV+ and RSV-deceased infants. The best solution stress value was 0.248. Some clustering of RSV+ decedents is visible to the bottom with some clustering of RSV-decedents to the top left. **B)** PCoA showing general clustering of RSV+ samples in the bottom right-hand corner without RSV-samples. However, otherwise there is substantial overlap between the two groups.

In the NMDS plot for genera, we observed clustering of RSV+ decedent samples to the bottom and RSV-decedent samples clustering to the upper left (Figure 3A). However, there was little spatial separation of clusters, and the stress value was high at 0.248. This indicates some differentiation between groups, but a high degree of overlap. NMDS for species also showed very little clustering by RSV status with a stress value of 0.262 (See Figure, Supplemental Digital Content 2A).

From the PCoA plot for genera, we see that the first two axes account for 38% of the variability in the data (Figure 2B). Some clustering is visible in the RSV+ samples, hedging away from the RSV-samples in the bottom right-hand corner with overlap between samples throughout the remainder. As with the NMDS analysis, signs of distinct clustering are largely absent. PCoA for species showed mostly overlap between RSV+ and RSV-samples with some individually clustered RSV+ samples in the bottom left (See Figure, Supplemental Digital Content 2B).

### Differential Abundance Analysis

Differential abundance analysis via the non-parametric Wilcoxon rank sum test was performed to compare differences in relative microbial composition between RSV+ and RSV-samples. From this test, we determined that *Gemella* (p=0.020), *Moraxella* (p=0.006) and *Staphylococcus* (p=0.018) all had marginally significant differences in median abundances between RSV+ and RSV-samples (See Table, Supplemental Digital Content 3).

We created box plots of genera abundances to visualize the inter-group differences. We observed that RSV+ decedents have higher abundances of *Gemella* and *Staphylococcus* and a lower abundance of *Moraxella* compared to RSV-decedents (Figure 4A). We also created a heatmap based on relative abundances. In the heatmap, we see subtle trends in abundance differences between the RSV+ and RSV-decedents consistent with our differential abundance results and box plots (See Figure 3A).

**Figure 4.**
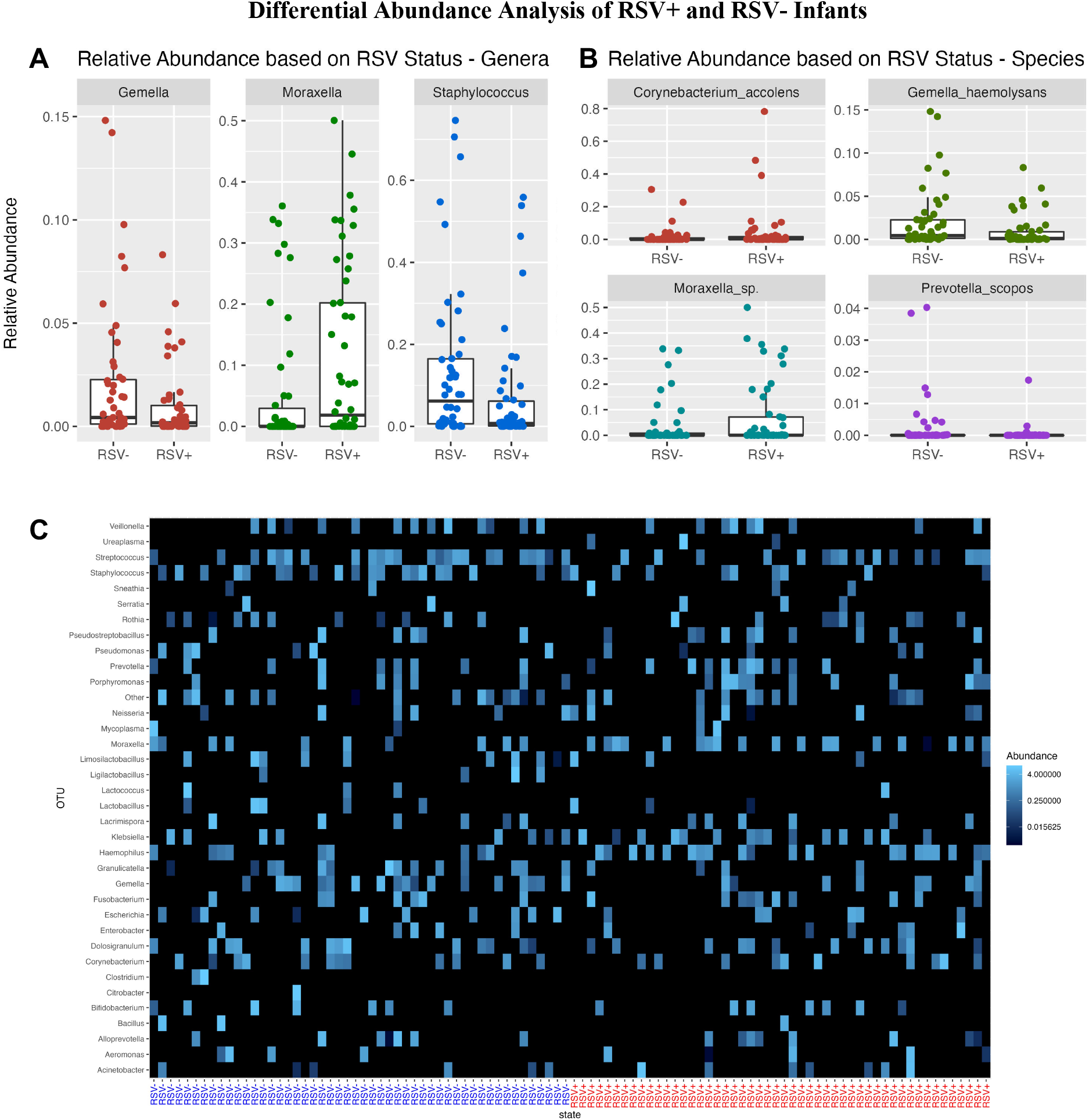
Box plots of relative abundance indicating significant **A)** intra-genera or **B)** intra-species differences in RSV+ and RSV-samples. **A)** RSV+ samples show lower abundances in the genera of *Gemella* (p=0.020) and *Staphylococcus* (p=0.018) and a higher abundance of *Moraxella* (p=0.006). **B)** RSV+ samples show higher abundances of the species *Moraxella sp*. (p=0.016) and lower abundances of *Gemella haemolysans* (p=0.006). Differences in *Corynebacterium accolens* (p=0.049) and *Prevotella scopos* (p=0.004) appear to be driven by a few outliers. **C)** Heatmap displaying the Bray-Curtis dissimilarity index for genera comparison between RSV+ and RSV-decedents. Higher abundances of *Haemophilus* and *Moraxella* are seen in RSV+ decedents. Lower abundances of *Gemella, Staphylococcus* and *Streptococcus* are present in RSV+ decedents.

Differential abundance analysis at the species level, which is possible using PathoScope2.0, indicated significant differences for *Corynebacterium accolens* (p=0.049), *Gemella haemolysans* (p=0.006), *Moraxella sp*. (p=0.016), and *Prevotella scopos* (p=0.004).^22–24^ The box plots in Figure 4B illustrate higher abundances of *Moraxella sp*. in RSV+ decedents and lower abundances of *Gemella haemolysans* when compared with RSV-decedents. Differences in *Corynebacterium accolens* (p=0.049) and *Prevotella scopos* appear to be driven by a few outliers. Heatmap visualization revealed similar visible differences (See Figure, Supplemental Digital Content 4).

### Pathway Analysis

Using PICRUSt2, we inferred pathway abundances for each sample based on the microbial abundances computed with PathoScope.^22–24,44^ We analyzed these abundances using a Wilcoxon rank sum test to determine which pathways were differentially present between RSV+ and RSV-decedents. This provided 20 different pathways; of these, 3 pathways were known to be correlated with *Haemophilus influenzae* (See Table, Supplemental Digital Content 5). Of the remaining 17 pathways, 10 were correlated with *E. Coli*, 4 correlated to a variety of plant species, and 3 correlated to other infectious diseases (*Mycobacterium tuberculosis* and various species of *Chlamydia*).

## DISCUSSION

Previous studies have shown that the microbial ecosystem in the nasopharynx, in which RSV acquisition occurs, may influence the host response to infection and certainly varies with disease severity.^2,8–13^ In contrast with prior studies, which focused mainly on infants who were newly diagnosed with RSV and were acutely ill, our study focused on deceased infants who died of a respiratory cause, namely RSV. This focuses our results on infants who had experienced the most extreme outcome of RSV infection, death. Previous studies have also shown that antibiotic use leads to decreased alpha diversity and dysbiosis in the NP microbiome.^14,15^ To avoid confounding by exposure to antibiotics, our analysis focused on comparing the microbiome profile of infants who died in the community. This includes infants who died in a facility with a stay less than 48 hours who would not be affected by nosocomial exposure due to limited time in the hospital.

We found no significant difference in genera or species richness or diversity in individual samples as measured by the Shannon and Simpson indexes. This indicates that the number of different species was roughly similar between groups but does not provide insight into whether the specific compositions differed. To explore how the specific compositions of the NP microbiome differed, we turned to examine beta diversity via Bray-Curtis analysis. NMDS and PCoA plots showed high degrees of overlap, but PERMANOVA indicated a significant difference between the two groups (p<0.001). To explore which microbes influenced the difference in beta diversity, we analyzed differences in the relative abundance of microbial taxa between conditions. RSV+ decedents had higher abundances of *Moraxella* (p=0.006) and lower relative abundances of *Gemella* (p=0.020) and *Staphylococcus* (p=0.018) when compared to RSV-decedents. We also noticed increased relative abundances of *Haemophilus* in RSV+ decedents, although these were not significant. We identified significant differential abundance in 3 pathways associated with *Haemophilus*. This confirms findings reported previously by Ederveen et al., Rosas-Salazar et al. and de Steenhuijsen Piters et al.^11–13^

Ederveen et al. reported that an overabundance of *Haemophilus* and *Achromobacter* microbes and a loss of microbes like *Veillonella* are associated with infants who had been hospitalized with RSV.^12^ They also indicated that infants who recover from RSV have higher *Moraxella* abundances. Additionally, Rosas-Salazar et al. observed that infants diagnosed with RSV had higher abundances of *Haemophilus, Moraxella*, and *Streptococcus* whereas healthy patients had high abundances of *Corynebacterium* and *Staphylococcus*.^11^ Also, de Steenhuijsen Piters et al. reported that in infants seen in a clinic or hospitalized for RSV, *H. influenzae* and *Streptococcus* are seen in higher abundance and correlated to disease severity.^13^ Our study found similar results in infants who died with RSV in *Haemophilus* and *Moraxella*. Differences in our study and those by these mentioned studies may be attributed to the fact their study participants received medical care in a clinical or hospital setting. While our infants represent the most severe disease status (resulting in death), none of the infants in our study were treated for more than 48 hours in a clinical setting before passing away and most of the cases in our study did not receive any exposure to a clinical setting in the course of disease. Differences in some of the microbes, such as *Streptococcus*, may be due to exposure to clinical settings and/or usage of antibiotics. Our results confirm that higher abundances of *Haemophilus* and *Moraxella* and decreases in *Staphylococcus* are correlated with severe RSV infection.

Our study had several key limitations. For one, we did not have complete data on how the infants in either population died. Despite the adjudication process (See Text, Supplemental Digital Content 1), there is a chance that the RSV+ group may have died from issues unrelated to RSV or respiratory disease which could skew our results. Compounding this, information about the final HIV status of the decedents is not available and HIV status of the mothers may not be accurately captured since mother’s HIV status was not always noted in medical charts or death records and was not inquired about during verbal autopsies. HIV exposure has been shown to cause NP microbiome dysbiosis and may affect our results.^45,46^ Furthermore, many of the decedents, particularly those who had died in a facility, may have been treated with antibiotics. While our focus on community deaths was intended to reduce the confounding effect of antibiotics, we could not exclude the possibility that some decedents were exposed to antibiotics without our knowledge. We also do not have information regarding use of antibiotics earlier in life which may also disrupt the nasopharyngeal microbiome.^15^ Additionally, verbal autopsies did not yield accurate information about antibiotic exposure. Lastly, the impact of RSV on microbial ecology may not necessarily be confined to the acute RSV infection period but could occur at some time removed from the initial infection. The cross-sectional nature of our data set, focusing on time point shortly after death, could not detect those delayed effects.

With those caveats, our observations are consistent with what has been identified in previous research by Ederveen et al., Rosas-Salazar et al. and de Steenhuijsen Piters et al.

## CONCLUSION

Our findings support those findings previously published and indicate that RSV may be correlated with higher abundances of *Moraxella* and *Haemophilus* and lower abundances of *Staphylococcus*. Our study also points to decreased abundances of *Gemella*.

Further research should be done to confirm these findings and specifically to determine if the change in microbiome proceeds RSV infection or is an effect of infection. Additional research looking at community-acquired RSV would also allow greater understanding of microbe differences without exposure to clinical settings or antibiotics. Better understanding may allow for better identification of at-risk infants and lead to better treatment and care.

## Supporting information

Supplemental Digital Content 2

Supplemental Digital Content 3

Supplemental Digital Content 4

Supplemental Digital Content 5

Supplemental Digital Content 6

Supplemental Digital Content 1

## Data Availability

Raw data is available on the sequence read archive, accession number PRJNA913857. All processed data and code used for data preprocessing, statistical analysis, and figure generation is available on GitHub (https://github.com/jessmcc22/ZPRIME_RSV).

https://www.ncbi.nlm.nih.gov/sra

## Author Contribution Statement

CJG and LM were the co-principal investigators for the ZPRIME study. WBM was the ZPRIME study statistician and provided all needed data from the ZPRIME study for our analysis. AI oversaw the lab that generated the microbiome samples from ZPRIME for our study. RP was the ZPRIME project manager and aided in collaboration between our teams. RL contributed to the implementation of the ZPRIME study, including with the adjudication process. WEJ oversaw all data and statistical analysis for the current study and provided recommendations on best practices. NK completed initial data exploration and analysis. JM revised and completed the data visualization, data analysis and statistical analysis. She also took the lead on writing the manuscript. AROM provided support for data and statistical analysis, template code for sequencing, animalcules and PICRUSt analysis, and provided critical feedback on the manuscript. JM, AROM, CJG, WBM, RP, RL and WEJ discussed the results, contributed to the ideas contained in the final manuscript, and helped prepare the manuscript. All authors reviewed the final manuscript.

## Data Access Statement

Raw data is available on the sequence read archive, accession number PRJNA913857. All processed data and code used for data preprocessing, statistical analysis, and figure generation is available on GitHub (https://github.com/jessmcc22/ZPRIME_RSV). Sample IDs are consistent across microbiome and demographic information on the sequence read achieve and GitHub. Full results for all analyses are also available on GitHub.

## Ethics Statement

The ZPRIME study, where our data originates, was approved by the ethical review boards at Boston University Medical Center and the University of Zambia. Written informed consent was obtained from the decedents’ next of kin or guardian.

## List of Supplemental Digital Content

Supplemental Digital Content 1. Text: Methods continued

Supplemental Digital Content 2. Figure: Species Level Bray-Curtis Dissimilarity Index

Supplemental Digital Content 3. Table: p-values and adjusted p-values for RSV Differential Abundance Analysis

Supplemental Digital Content 4. Figure: Species Level Differential Abundance Heatmap

Supplemental Digital Content 5. Table: PICRUSt Pathway Analysis Significant Results

Supplemental Digital Content 6. Table: STORMS Checklist

