## Supplemental Digital Content 2 for "Post-mortem Nasopharyngeal Microbiome Analysis of Zambian Infants with and without Respiratory Syncytial Virus Disease: A Nested Case Control Study"

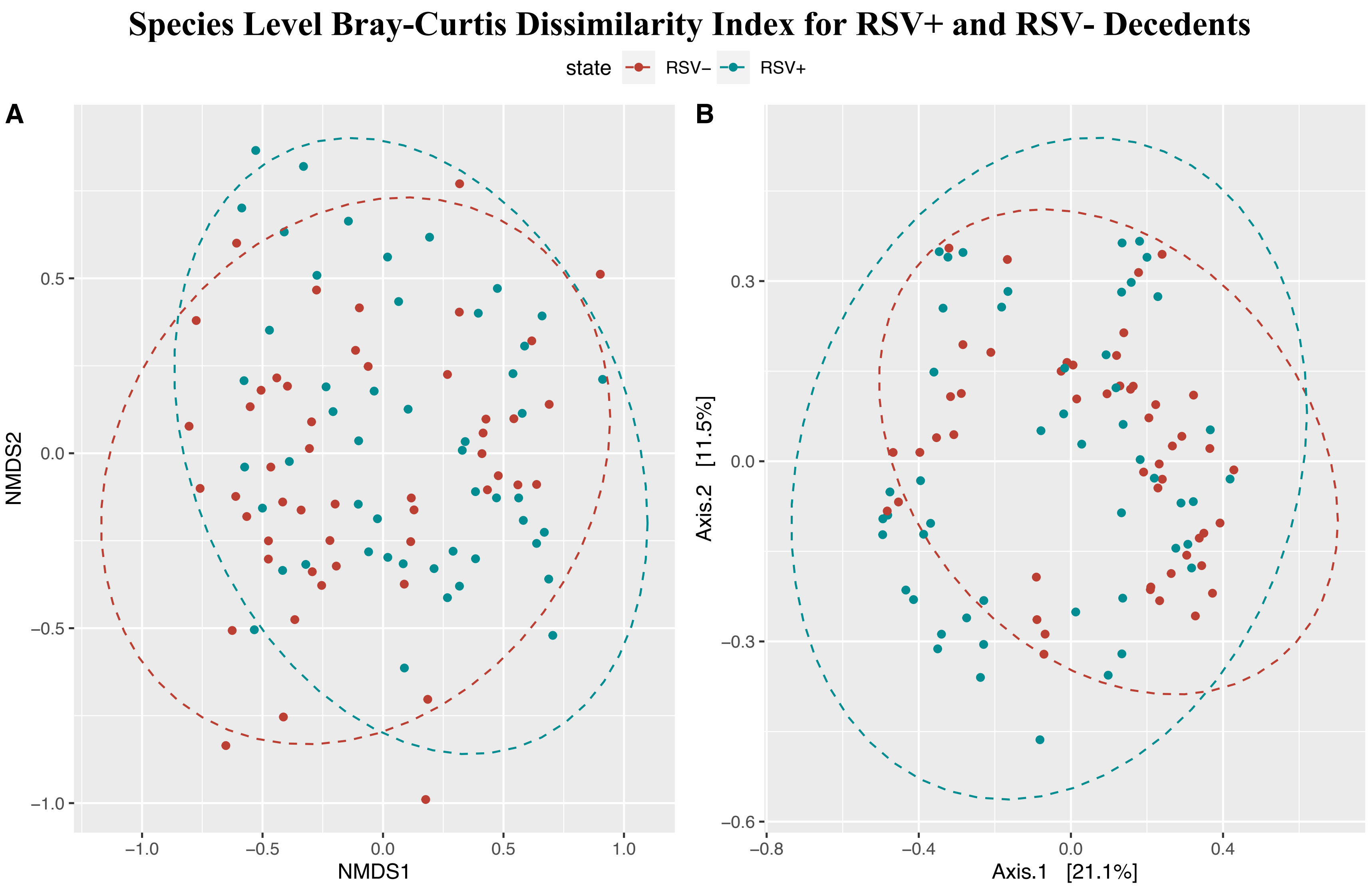


**Figure, Supplemental Digital Content 3** Species analyses displaying the Bray Curtis dissimilarity index between RSV+ and RSV- infants. **A)** NMDS overlap between RSV+ and RSV- samples with a stress value of 0.262 showing very little distinct clustering. **B)** PCoA indicating some unique clustering of RSV+ in bottom left-hand corner.
