## Supplemental Digital Content 3 for "Post-mortem Nasopharyngeal Microbiome Analysis of Zambian Infants with and without Respiratory Syncytial Virus Disease: A Nested Case Control Study"

**p-values and Adjusted p-values for Differential Abundance Analysis at Genus Level**

| Genera | p-value | Adjusted p-value |
|---|---|---|
| *Acinetobacter* | 0.885 | 0.969 |
| *Aeromonas* | 0.447 | 0.838 |
| *Alloprevotella* | 0.829 | 0.969 |
| *Bacillus* | 0.747 | 0.969 |
| *Bifidobacterium* | 0.889 | 0.969 |
| *Citrobacter* | 0.973 | 0.978 |
| *Clostridium* | 0.978 | 0.978 |
| *Corynebacterium* | 0.871 | 0.969 |
| *Dolosigranulum* | 0.267 | 0.705 |
| *Enterobacter* | 0.124 | 0.446 |
| *Escherichia* | 0.117 | 0.446 |
| *Fusobacterium* | 0.274 | 0.705 |
| *Gemella* | 0.02** | 0.24 |
| *Granulicatella* | 0.08 | 0.446 |
| *Haemophilus* | 0.059 | 0.425 |
| *Klebsiella* | 0.393 | 0.838 |
| *Lacrimispora* | 0.565 | 0.838 |
| *Lactobacillus* | 0.915 | 0.969 |
| *Lactococcus* | 0.519 | 0.838 |
| *Ligilactobacillus* | 0.141 | 0.461 |
| *Limosilactobacillus* | 0.582 | 0.838 |
| *Moraxella* | 0.006*** | 0.216 |
| *Mycoplasma* | 0.426 | 0.838 |
| *Neisseria* | 0.508 | 0.838 |
| *Other* | 0.357 | 0.838 |
| *Porphyromonas* | 0.11 | 0.446 |
| *Prevotella* | 0.551 | 0.838 |
| *Pseudomonas* | 0.707 | 0.969 |
| *Pseudostreptobacillus* | 0.877 | 0.969 |
| *Rothia* | 0.268 | 0.705 |
| *Serratia* | 0.486 | 0.838 |
| *Sneathia* | 0.547 | 0.838 |
| *Staphylococcus* | 0.018** | 0.24 |
| *Streptococcus* | 0.051 | 0.425 |
| *Ureaplasma* | 0.793 | 0.969 |
| *Veillonella* | 0.099 | 0.446 |

**Table, Supplemental Digital Content 2.** Table of Wilcoxon rank sum test p-values testing RSV status differences with a Benjamini-Hochberg multiple testing adjustment. Significant values are marked with *** $p < 0.01$, ** $p < 0.03$, * $p < 0.05$.
