## Supplemental Digital Content 4 for "Post-mortem Nasopharyngeal Microbiome Analysis of Zambian Infants with and without Respiratory Syncytial Virus Disease: A Nested Case Control Study"

### Species Level Heatmap of Differential Abundance

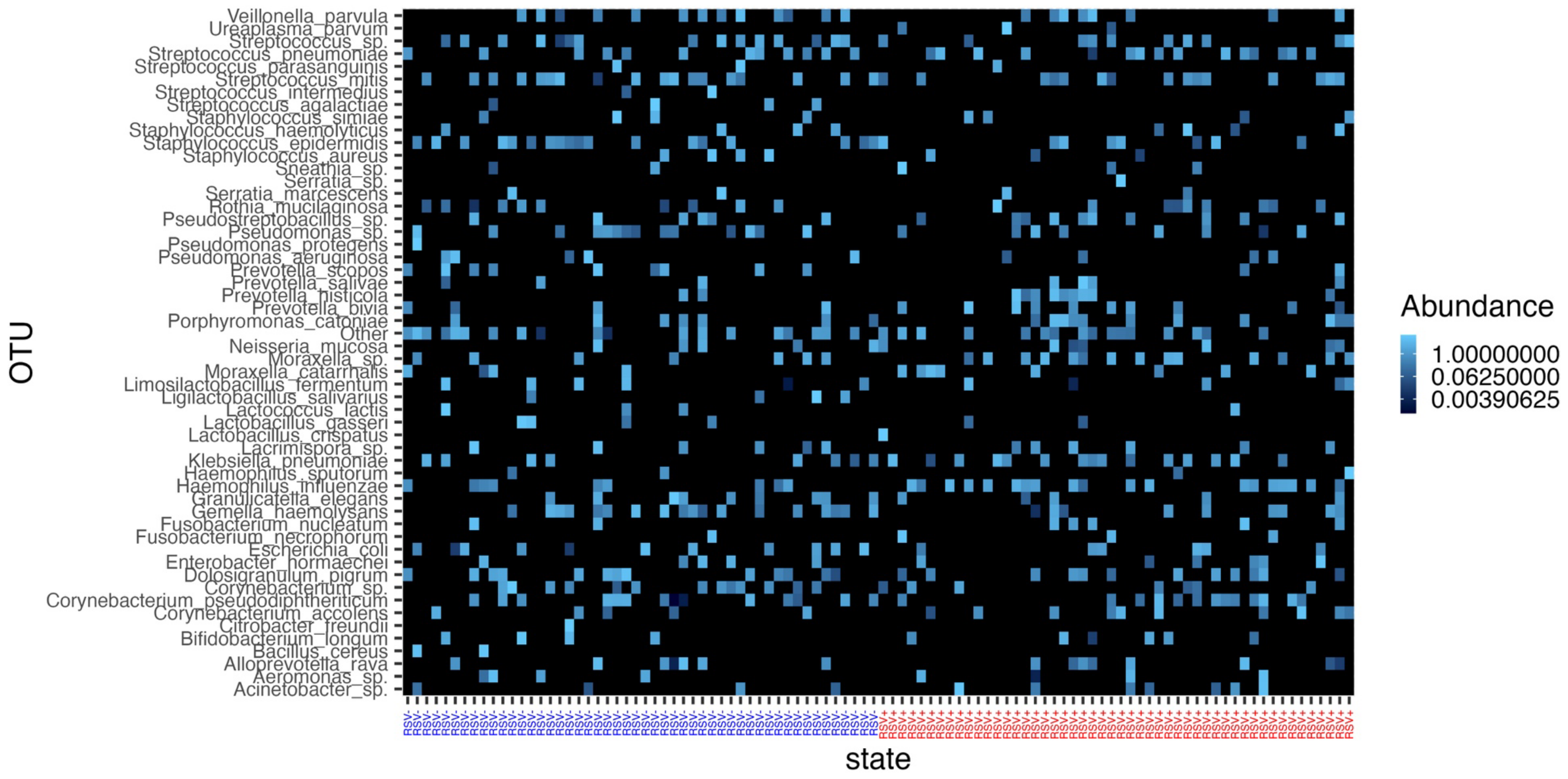


**Figure, Supplemental Digital Content 4.** Heatmap showing species differential abundance comparison. Note differences for *Corynebacterium accolens*, *Gemella haemolysans, Moraxella sp*., and *Prevotella scopos*.
