## Supplemental Digital Content 5 for "Post-mortem Nasopharyngeal Microbiome Analysis of Zambian Infants with and without Respiratory Syncytial Virus Disease: A Nested Case Control Study"

### PICRUSt Pathway Abundance Analysis Significant Results

| Pathway | Taxa w/pathway (per MetaCyc) | Adjusted p-value |
|---|---|---|
| 6-hydroxymethyl-dihydropterin diphosphate biosynthesis III (Chlamydia) | *Chlamydia trachomatis* | 0.029 |
| 8-amino-7-oxononanoate biosynthesis I | *Candidatus Nitrososphaera gargensis Ga9.2*<br>*Escherichia coli K-12 substr. MG1655*<br>*Francisella tularensis novicida U112*<br>***Haemophilus influenzae Rd KW20***<br>***Helicobacter pylori 26695***<br>***Mycobacterium tuberculosis H37Rv***<br>***Prochlorococcus marinus MIT 9211*** | 0.007 |
| biotin biosynthesis I | *Escherichia coli K-12 substr. MG1655*<br>*Francisella tularensis novicida U112*<br>***Haemophilus influenzae Rd KW20***<br>***Helicobacter pylori 26695***<br>***Prochlorococcus marinus MIT 9211*** | 0.007 |
| CMP-3-deoxy-D-manno-octulosonate biosynthesis I | *Arabidopsis thaliana col*<br>*Escherichia coli BL21(DE3)*<br>*Escherichia coli K-12 substr. MG1655*<br>*Zea mays* | 0.003 |
| flavin biosynthesis I (bacteria and plants) | *Arabidopsis thaliana col*<br>*Bacillus subtilis*<br>*Escherichia coli K-12 substr. MG1655* | 0.029 |
| fucose degradation | *Escherichia coli K-12 substr. MG1655* | 0.045 |
| Kdo transfer to lipid IVA III (Chlamydia) | *Chlamydia pneumoniae*<br>*Chlamydia psittaci*<br>*Chlamydia psittaci 6BC* | 0.003 |
| L-lysine biosynthesis I | *Arthrobacter globiformis*<br>*Azotobacter vinelandii*<br>*Bordetella pertussis*<br>*Corynebacterium glutamicum*<br>*Corynebacterium glutamicum ATCC 13032*<br>*Escherichia coli K-12 substr. MG1655*<br>***Haemophilus influenzae***<br>***Haemophilus influenzae Rd KW20***<br>*Helicobacter pylori*<br>*Mycobacterium tuberculosis*<br>*Mycobacterium tuberculosis H37Rv*<br>*Rhodospirillum rubrum* | 0.034 |
| L-lysine biosynthesis VI | *Arabidopsis thaliana col*<br>*Chlamydia trachomatis*<br>*Glycine max*<br>*Methanocaldococcus jannaschii*<br>*Methanothermobacter thermautotrophicus*<br>*Nicotiana tabacum*<br>*Synechocystis*<br>*Zea mays* | 0.026 |
| lipid IVA biosynthesis | *Caulobacter vibrioides NA1000*<br>*Escherichia coli K-12 substr. MG1655* | 0.003 |
| methylerythritol phosphate pathway I | *Brucella abortus*<br>*Escherichia coli K-12 substr. MG1655* | 0.005 |
| methylerythritol phosphate pathway II | *Arabidopsis thaliana col*<br>*Botryococcus braunii*<br>*Croton stellatopilosus*<br>*Plasmodium falciparum*<br>*Plasmodium falciparum HB3*<br>*Solanum lycopersicum*<br>*Thermosynechococcus vestitus BP-1* | 0.005 |

| | | |
|---|---|---|
| mycolate biosynthesis | *Mycobacterium tuberculosis H37Rv* | 0.023 |
| NAD salvage pathway II | *Escherichia coli K-12 substr. MG1655*<br>*Salmonella enterica enterica serovar Typhimurium* | 0.009 |
| oleate biosynthesis IV (anaerobic) | *Aerococcus viridans*<br>*Cereibacter sphaeroides 2.4.1*<br>*Clostridium beijerinckii* | 0.023 |
| palmitate biosynthesis II (bacteria and plants) | *Arabidopsis thaliana col*<br>*Brassica napus*<br>*Escherichia coli K-12 substr. MG1655*<br>*Spinacia oleracea* | 0.025 |
| palmitoleate biosynthesis I (from (5Z)-dodec-5-enoate) | *Escherichia coli K-12 substr. MG1655*<br>*Helicobacter pylori 26695* | 0.003 |
| stearate biosynthesis II (bacteria and plants) | *Arabidopsis thaliana col*<br>*Brassica napus*<br>*Spinacia oleracea* | 0.012 |
| superpathway of (Kdo)2-lipid A biosynthesis | *Escherichia coli K-12 substr. MG1655* | 0.028 |
| superpathway of fatty acid biosynthesis initiation (E. coli) | *Escherichia coli K-12 substr. MG1655* | 0.030 |

**Table, Supplemental Digital Content 5.** PICRUSt2 inferred pathways and associated species. These pathway abundances are all significantly different between RSV+ and RSV- samples. Species identified as being differentially expressed in RSV+ and RSV- decedents in our differential abundance analysis are bolded.
