## Supplemental Digital Content 1 for "Post-mortem Nasopharyngeal Microbiome Analysis of Zambian Infants with and without Respiratory Syncytial Virus Disease: A Nested Case Control Study"

**Text, Supplemental Digital Content 1.**

### Study Population and Study Design

The ZPRIME study systematically enrolled infants that died at ages ranging from four days to six months in Lusaka, Zambia between 2017 and 2020, omitting a 6-week window in 2020 due to restrictions as a result of COVID-19. Decedents were enrolled within 48 hours of death and, for our study, were either brought in dead (BID) or died in a facility within 48 hours of hospitalization (denoted as an early facility death). Early facility deaths were included as community deaths since these decedents would likely not be affected by nosocomial exposure due to limited time in the hospital. Study activities were concentrated at the University Teaching Hospital and associated morgue in Lusaka. For early facility deaths, these data were abstracted from medical chart records and official death certificates. For BID infants, we interviewed the decedents' next of kin and collected a verbal autopsy that focused on identifying whether the infant had a respiratory illness as the cause of death. Selection bias may be present since our study only focused on deaths in the morgue associated with the University Teaching Hospital where over 80% of all deceased individuals in Lusaka pass through. Some community deaths will not be captured in our study population and thus cannot be selected for participation.

Three study clinicians reviewed the verbal autopsy or clinical information available for each death to determine if the cause of death was respiratory, non-respiratory, or uncertain. Discordant adjudications were discussed among the three clinicians and either resolved or labelled as uncertain if agreement could not be reached. The clinicians were blinded to the laboratory results during this process.

From these samples, we selected a subset of community deaths for our analysis. We limited our study to infants that were either BID or early facility deaths marked as RSV+ with a PCR Ct<35. We upheld a more stringent RSV cut off in our criteria to exclude incorrect RSV diagnoses within our study, thereby increasing specificity. Additionally, we considered infants who died within 48 hours of admittance to a facility as a community death as it is unlikely that their microbiome would have been affected by nosocomial exposure. However, antibiotic usage was not known for any of the deceased infants included in this study.

### Sample Collection, Processing, and Storage

All NP samples were collected from decedents at the time of enrollment into ZPRIME. NP samples were obtained using flocked-tipped nylon swabs (Copan Diagnostics, Murrieta CA) sized appropriately for infants. Two swabs were used per decedent. Each swab was inserted into either the right or left nostril until contacting the posterior nasopharynx, then rotated 180 degrees to the left and then to the right. The swabs were then placed in 3mL of universal transport media at 2–8°C, put on ice, and transferred to an onsite lab on the same campus (University Teaching Hospital in Lusaka, Zambia), where they were aliquoted and stored at -80°C until DNA extraction. DNA was extracted using the NucliSENS EasyMagG system (bioMerieux, Marcy, l'Etoile, France,) and was stored at the University Teaching Hospital at -80°C. Extracted DNA was stored at the lab located at the University Teaching Hospital in Lusaka at -80°C. Sample collection, processing and storage was previously described in Gill et al.[1]

### 16S rDNA Amplification and Sequencing

16S ribosomal DNA was amplified using PCR with primers specific to the V3–V4 region. PCR products and negative controls were visualized, the indexed libraries were purified and quantified, and sequenced as previously described in Lapidot et al, including control for contamination.[2,3] Samples were sequenced on the Illumina MiSeq sequencing platform at the National Institute for Communicable Diseases, Sequencing Core Facility (South Africa) following standard Illumina sequencing protocols.

### Data Processing

We used FastQC v0.11.9 to assess the quality of reads; our multiQC report is available on Github (https://github.com/jessmcc22/ZPRIME_RSV/blob/main/multiqc_report.html).[4] All reads (both forward and reverse) had a mean Phred score > 30 (> 99.9% accuracy), indicating a good sequence quality for all the samples included in this analysis. Illumina adapters were trimmed and low quality sequences were removed using Trimmomatic v.0.39 with the following parameters: SLIDINGWINDOW: 4:20, LEADING:3, TRAILING:3, and MINLEN:36.[5] We used PathoScope 2.0 with its default parameters to quantify the proportions of reads from individual microbial strains present in the metagenomic sequencing data.[6,7] We used Refseq bacterial and viral genomes which were downloaded on November 2, 2018 as PathoScope reference libraries.[8] We have made these available in our GitHub repository (https://github.com/jessmcc22/ZPRIME_RSV#readme). Analysis was completed at both the genus and species level, which has been shown to be possible using Pathoscope.[9] PICRUSt2 was used to infer pathway abundances based on the PathoScope output.[10–15]

All raw data, processed data, and code used for data preprocessing, statistical analysis, and figure generation is available on GitHub (https://github.com/jessmcc22/ZPRIME_RSV). Raw data is also available on the sequence read archive, accession number PRJNA913857. All data and statistical analysis were completed in R version 4.2.0, primarily using packages ALDEx2 v. 1.28.1, animalcules 1.12.0, ggplot2 3.3.6, ggpubr 0.4.0, paletteer 1.5.0, Phyloseq 1.40.0, tidyverse 1.3.1 and vegan 2.6-2.[16–23] Animalcules was used for initial exploratory visualization of the data and to create a Multi-Assay Experiment object from the raw data reads. From this object, a Phyloseq object was created and this Phyloseq object was used for statistical analysis moving forward, including creating the figures in this paper. All other packages were used for data manipulation, visualization, and statistical testing.
